# Estimating the Global Infection Fatality Rate of COVID-19

**DOI:** 10.1101/2020.05.11.20098780

**Authors:** Richard E. Grewelle, Giulio A. De Leo

**Affiliations:** Dept. of Biology, Stanford University, 371 Serra Mall, Stanford, CA; Hopkins Marine Station, 120 Oceanview Blvd., Pacific Grove, CA

**Author notes:** Corresponding author: Email address (Richard E. Grewelle).

**Keywords:** asymptomatic, seroprevalence, pandemic, testing capacity

## Abstract

COVID-19 has become a global pandemic, resulting in nearly three hundred thousand deaths distributed heterogeneously across countries. Estimating the infection fatality rate (IFR) has been elusive due to the presence of asymptomatic or mildly symptomatic infections and lack of testing capacity. We analyze global data to derive the IFR of COVID-19. Estimates of COVID-19 IFR in each country or locality differ due to variable sampling regimes, demographics, and healthcare resources. We present a novel statistical approach based on sampling effort and the reported case fatality rate of each country. The asymptote of this function gives the global IFR. Applying this asymptotic estimator to cumulative COVID-19 data from 139 countries reveals a global IFR of 1.04% (CI: 0.77%,1.38%). Deviation of countries’ reported CFR from the estimator does not correlate with demography or per capita GDP, suggesting variation is due to differing testing regimes or reporting guidelines by country. Estimates of IFR through seroprevalence studies and point estimates from case studies or sub-sampled populations are limited by sample coverage and cannot inform a global IFR, as mortality is known to vary dramatically by age and treatment availability. Our estimated IFR aligns with many previous estimates and is the first attempt at a global estimate of COVID-19 IFR.

## Introduction

SARS-COV-2 emerged from Wuhan, China at the end of 2019 and has become a global pandemic with over four million confirmed infections and nearly three-hundred thousand associated deaths [1, 2, 3, 4, 5]. Now spreading in all continents but Antarctica, the disease associated with this novel virus (COVID-19) is a risk to people of all nations [6]. Yet, this risk is known to vary widely with access to healthcare and demographic traits [3, 4, 7]. Although the extent of sub-lethal impacts of the virus are still not known, deaths attributed to infection are of great concern and are used as a measure of disease burden, being a more reliable metric than the number of confirmed infections as the latter is notoriously dependent on testing capacity [8].

Counter-measures to inhibit the transmission of the virus are in place in most countries, and economic decline has resulted from the closure of businesses and travel restrictions [9, 10]. With pressure to re-open economies to reduce job losses and downstream repercussions, leaders of countries and localities are weighing the health risks of COVID-19 with economic risks of control measures [11]. The linchpin for policymakers is often the risk of mortality upon infection. Reported data from countries reveals a large spread in case fatality rate (CFR) from 0.5-15% [12]. This rate is given by the number of deaths per confirmed infection. The vast difference in the reported CFR by country is primarily a product of testing availability [13, 14]. In the early phases of the outbreak and in countries where testing kits were limited, only hospitalized patients with advanced COVID-19 symptoms have been tested. Therefore, the CFR is an inflated estimate of the infection fatality rate (IFR) because many infections in the population are unidentified [15, 16]. This inflation is intrinsic for reported data across all countries, though it is much smaller for countries that have greater testing capacity such that individuals with mild or no symptoms are tested.

To understand how dramatic this inflation is, an increasing number of serological studies has contrasted the number of active infections confirmed with PCR-based tests with the number of individuals with a detectable SARS-COV-2 immune response in sampled populations [17]. Universally, seroprevalence surveys confirm the presence of unreported infections, though the extent of the unreported infections and associated adjustment to IFR is debated due to the non-random nature of population sampling, low sample size, and questioned specificity of serological assays. Case studies reporting IFR from whole population sampling, such as the Diamond Princess cruise where all patrons were tested and the calculated IFR is near 2% are not necessarily representative of global demographics, incomes, or healthcare availability [7, 18]. In the absence of high resolution information of case demographics, seroprevalence, or standardization of healthcare and underlying risk factors to COVID-19 morbidity and mortality, there exists no method to assess the global IFR. A robust estimate is essential to inform policy and interventions [8, 19]. We provide a new statistical approach to estimate the global IFR using reported data, despite variability in reporting and risk factors. Differences in reported country-level CFR from the estimator are compared to national demographic and economic profiles to determine whether the reported CFR reflects known risk factors for COVID-19 mortality.

## Methods

Reporting standards and testing procedures vary widely across countries, preventing reliable interpretation of reported CFR. High CFR are generally associated with restricted access to testing due to low test availability, such that only individuals with marked COVID-19 symptoms – typically those of older age or with pre-existing conditions, and thus at higher risk of death from COVID-19 – receive testing [9, 13, 14, 20]. Greater testing capacity allows individuals with mild symptoms or those with suspected exposure to receive testing. Figure 1 illustrates how this allows for a broader net of sampling in a population, giving a closer estimate of the number of total infections in the population.

**Figure 1:**
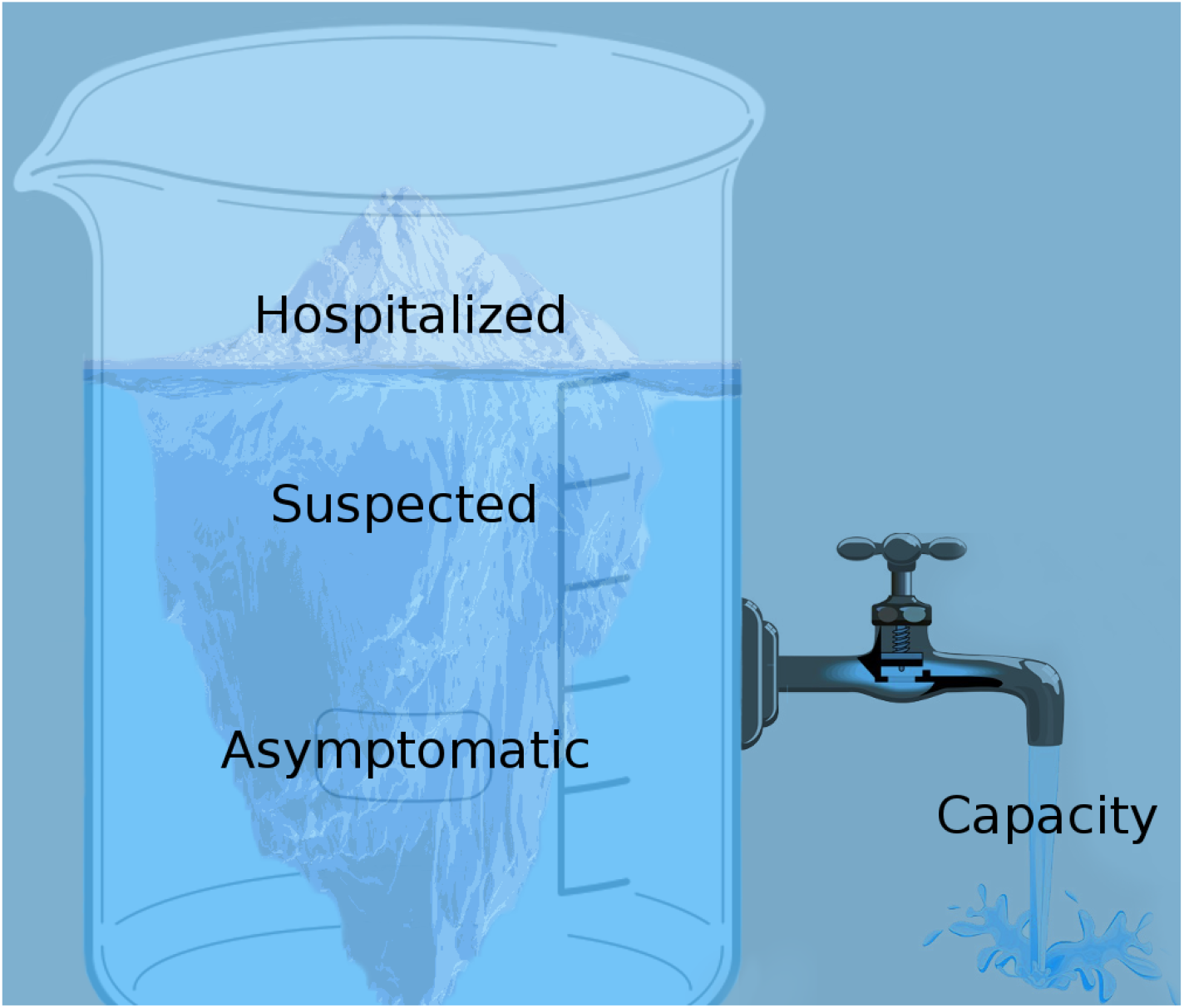
Increasing testing capacity uncovers a broader group of individuals, reducing estimates of the CFR for each country. Full capacity allows for estimation of the IFR.

As testing captures a greater proportion of the population, the estimated CFR better approximates the IFR because the individuals with mild or no symptoms and more likely to recover are included in the confirmed infections. With increasing testing capacity, the sample prevalence decreases. Testing capacity is relative to the case load in the country. A country with 1000 true infections requires more tests than a country with 100 true infections for similar case estimates. Because the true case load of COVID-19 is unknown in each country, we estimate testing capacity, *C*, as tests performed per positive case. This estimate provides a metric of the availability of tests relative to the case load, indicating relative testing capacity.

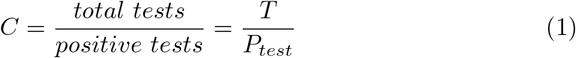

Capacity ranges from 1, where all tests are positive, to ∞, where all tests are negative. As capacity increases towards ∞ the estimate of IFR improves assuming testing targets people at risk of infection. This argument can be demonstrated by taking the inverse of *C*:

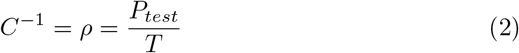

*ρ* is the sample prevalence. It is true that as the number of total tests increases and approaches a randomized sample of the whole population, the number of positive tests better estimates the active true case load in the population, *P_pop_*. The relation between *ρ* and the estimate of the true case load differs by testing regimes and cannot be fully known, but if testing prioritizes individuals with symptoms or at risk of infection, convergence of the known positive, *P_test_*, to the true positive, *P_pop_*, approximates an exponential decay with decay constant *k*, as shown in SI 1.

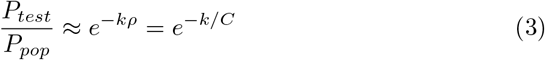

Despite differences in testing regimes in each country, the approximation in equation 3 becomes closer to exact as testing capacity increases. As *C* → ∞, *P_test_ → P_pop_*. From equation 3 the following is true:

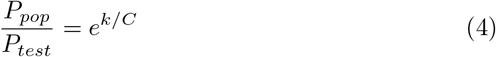

Log transformation yields:

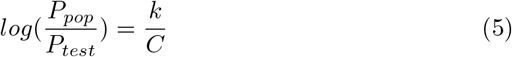

Assuming COVID-19 attributed deaths are more reliably estimated than COVID-19 cases, we can derive a relation to the true IFR from the reported CFR.

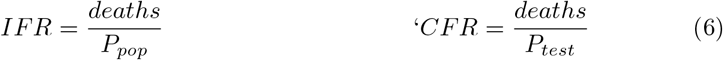

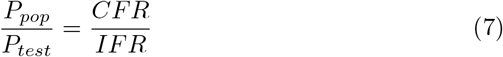

Substitution of this relation in equation 5 gives

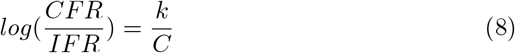

which is equivalent to

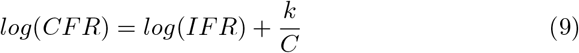

Equation 9 reveals that the y-intercept of a linear regression of *C*^-1^ = *ρ* and log transformed CFR can be exponentiated to yield the true IFR. We compile a comprehensive list of all countries that have recorded COVID-19 associated deaths as of April 21, 2020. After calculating the CFR, *P_test_, ρ*, and *C* for each country using cumulative cases, deaths, and tests performed, a weighted linear regression was performed according to equation 10:

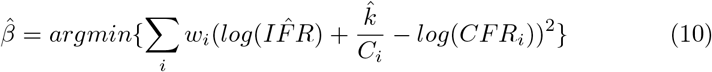

Countries are inverse variance weighted (*w_i_*); in the absence of variance attributed to testing regimes for each country, we assume homoscedasticity and variance of each estimate is inversely proportional to the number of tests performed (SI 2). The right panel is the plot of testing capacity and CFR with the regression achieved by equation 9 overlayed on the plot. The exponential of the y-intercept (at *ρ* = 0) in the regression corresponds to the asymptote as *C* → ∞ in panel B.

The variance-covariance matrix (**V**) of the parameter estimates, 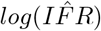 and 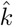, is calculated as

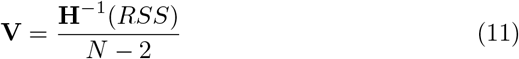

where **H**^−1^ is the inverted numerically estimated Hessian, *RSS* is the weighted residual sum of squares minimized in equation 10, and *N* is the number of countries. Confidence interval calculations are given in SI 4. A t-test for unequal variances was performed for each country, comparing its *log*(*CFR*) to the 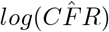 predicted by the linear regression for the given *ρ*. Low p-values indicate a low reported CFR relative to the global mean for a given *ρ*. P-values that are exceptionally low (p < 0.05) or exceptionally high (p> 0.95) indicate significant deviation from the expected global CFR. Through linear regression we investigated whether these deviations might be explained by per capita GDP or the fraction of the populace 65 years and older, as these national metrics could correlate with the national IFR. Additionally, these two variables were evaluated as possible predictors for a country’s reported CFR, independent of the relationship to testing capacity. Evaluation occurred through model selection via AIC in a GLMM framework using R Stan.

## Results

The transformation of the data from 139 countries according to equation 9 yields a linear relationship between sample prevalence (*ρ*) and the logarithm of reported CFR for each country.

The weighted regression produces estimates for the y-intercept at 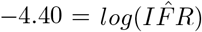 and slope at 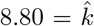. *R*^2^ = 0.5524, signaling that the relationship described between testing capacity and reported CFR explains over 55 % of the variation in the reported data. Exponentiating the y-intercept estimate yields a global IFR of 0.0123, or 1.23%. This corresponds to the asymptote seen in panel B of Figure 2 as *C* → ∞. 95% confidence intervals for *IFR* and *k* are (0.0096, 0.0157) and (8.1, 9.5), respectively. Because recent evidence suggests up to 15% false negative rate for rapid SARS-COV-2 PCR based tests, we adjust the calculations of capacity and CFR for each country and perform and adjusted regression which yields an intercept at 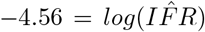. The adjusted IFR accounting for false negative tests is 1.04%. Due to the uncertainty associated with the 15% false negative approximation, we retain the larger confidence intervals of the unadjusted estimate to give a 95% confidence interval of (0.0077, 0.0138). These results support a global IFR between 0.77% and 1.38%. This estimate implicitly accounts for asymptomatic cases and country-level differences in demography and testing regimes, and as such, it represents a global average.

**Figure 2:**
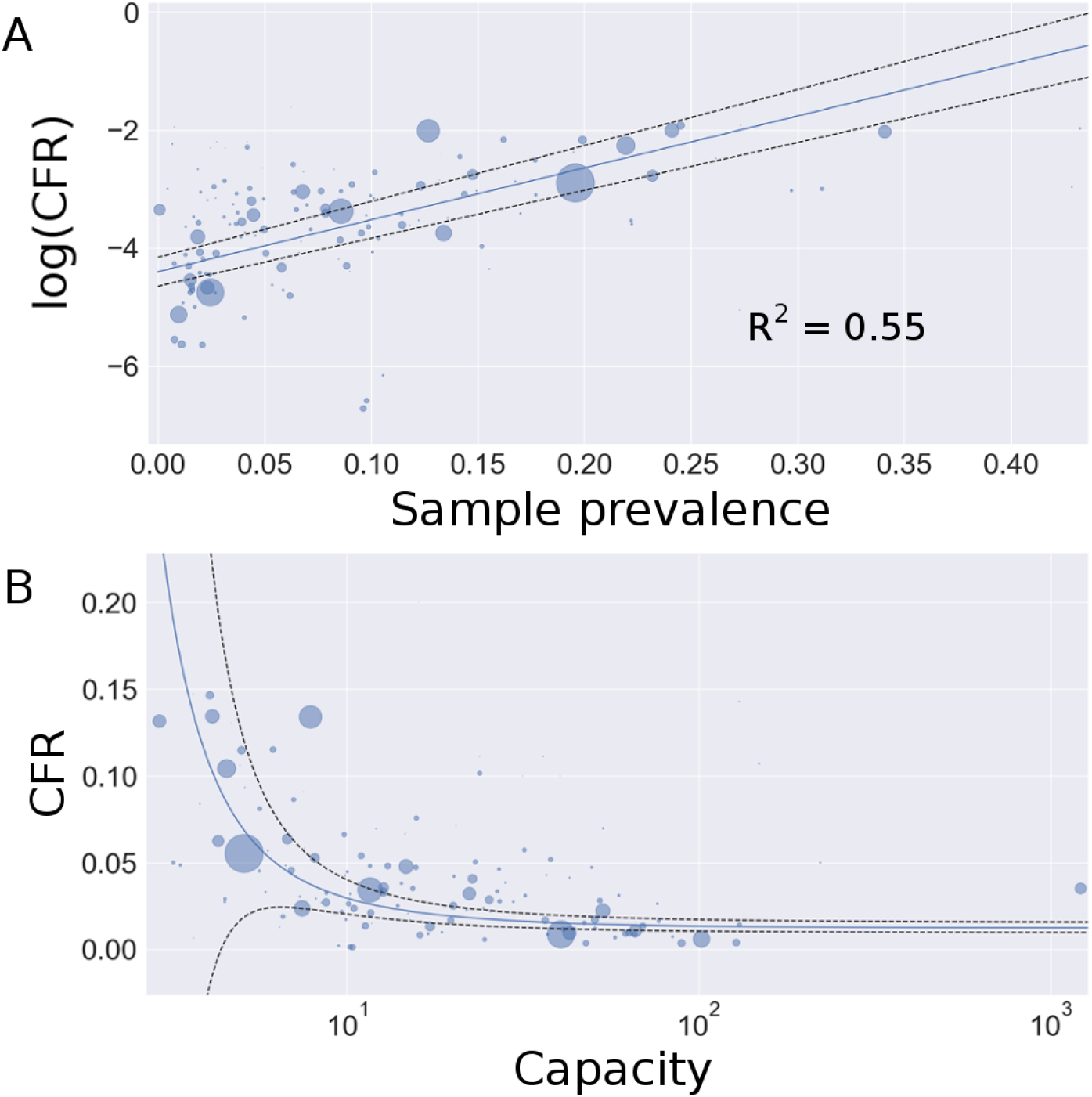
Weighted regression of reported national CFR and testing capacity. Circle area is proportional to each country’s weight. Panel A: linear regression defined by equation 10. The intercept corresponds to the logarithm of the estimated global IFR. Panel B: data plotted with the exponentiated regression line. The x-axis is log-scale for visualization (see Figure S1 for linear scale). The asymptote as *C* → ∞ corresponds to the estimated global IFR. Black dashed lines represent 95 % confidence intervals.

Reported here is a function relating testing capacity to the CFR estimate, and deviation from that function by a country may convey information of the underlying IFR. Placement above the regression line in panel B of Figure 2 could indicate higher national true IFR than the expected global average. Among the most reliable factors to predict IFR from COVID-19 are income and the age structure of the population, particularly the proportion of individuals 65 and older [7, 21]. We tested whether the reported CFR for each country was higher than the expected global mean at the corresponding testing capacity. p-values *<* 0.05 denote a lower CFR than the global mean, while p-values *>* 0.95 denote a higher CFR than the global mean. Figure 3 plots these p-values with World Bank data (2018) on country level per capita GDP and the percent of the populace 65 years or older [22, 23].

**Figure 3:**
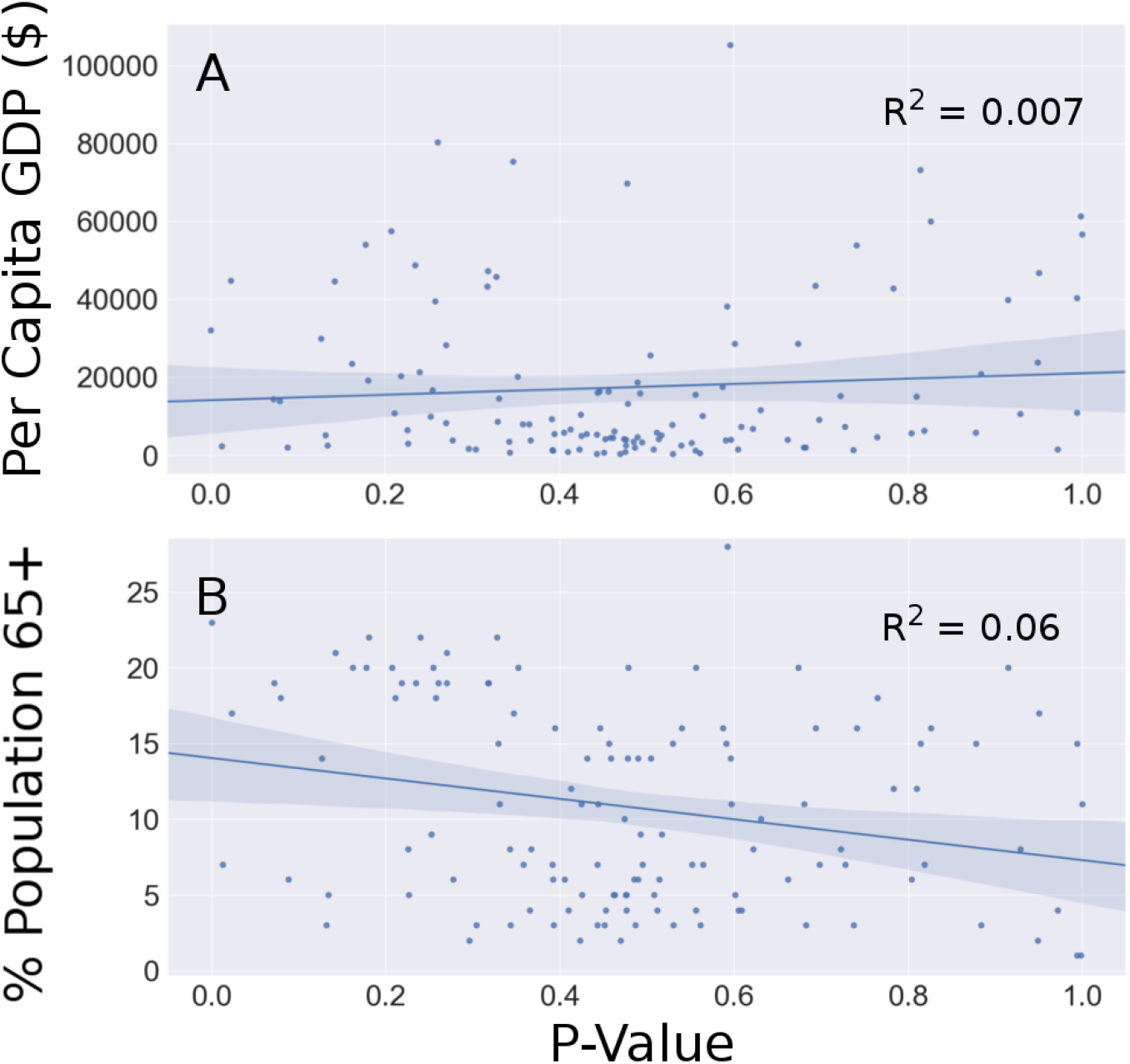
Relationships between P-values of the deviation of each country’s reported CFR from the global mean and risk factors for mortality from COVID-19. Larger p-values correspond to lower reported IFR relative to the global mean. Panel A: per capita GDP serves as a proxy for income, with lower income being a known risk for COVID-19 mortality. Per capita GDP is not predictive of differences in national IFR relative to the global mean. Panel B: the proportion of individuals 65 and older in the national population is also not predictive of national deviation from global CFR estimates.

Linear regressions reveal slight trends in favor of a lower CFR with increasing GDP and higher CFR with increasing age. These trends are marginal, and correlation between deviation from the global estimate and these factors is not substantiated. Rather, deviation is likely due to differences in COVID-19 reporting practices, testing, and stage of the epidemic in each country. Table 1 gives a list of the countries deviating significantly from the predicted CFR (all countries listed in SI 5). Countries with high p-values have a lower than expected CFR, which could indicate more conservative guidelines for attributing deaths to COVID-19. Conversely, countries with low p-values have a higher than expected CFR, which could occur via liberal guidelines for attributing deaths to COVID-19. Alternatively, extreme p-values could occur when testing does not prioritize at-risk groups (e.g. occurs randomly) or when testing reporting differs substantially from other countries, barring influence from risk factors like age, income, or access to healthcare.

**Table 1:** Countries that deviate substantially from the global trend in reported IFR. Countries are ordered by p-value (low to high). The dashed line separates countries with significantly higher (*α* < 0.05) reported CFR from countries with significantly lower (*α* > 0.95) reported CFR.

| Country | P-value |
| --- | --- |
| Italy | 2.27E-6 |
| Venezuela | 0.013 |
| Canada | 0.023 |
| ----- |  |
| Hong Kong | 0.950 |
| Uzbekistan | 0.972 |
| Russia | 0.994 |
| UAE | 0.994 |
| Qatar | 0.998 |
| Singapore | 0.999 |

National per capita GDP and the percent of the populace 65 years and older may also improve the estimate of a country’s reported CFR over that provided by testing capacity alone. Generalized linear mixed models including these two additional predictor variables were compared via AIC to reveal that the model including all 3 variables, *ρ*, per capita GDP, and % 65+, out-performed other models, including the model with *ρ* alone (SI 8). Results of the model comparison are reported in Table 2. Alone, neither GDP nor age were strong predictors of a nation’s reported CFR. The weak relationships aligned well with the relationships shown in Figure 3, where CFR increases with an aging populace and decreases with income. Due to the weakness of prediction, these models were not included in model comparison. Four models were compared, expanding on *ρ*. Age and GDP marginally improved the model fit, suggesting a small but non-significant influence of these factors in a country’s CFR. The relationship between age and GDP on a country’s IFR are unclear from these results because the most significant predictor of CFR is testing capacity.

**Table 2:** Ranked model comparison demonstrating improved model performance with inclusion of per capita GDP and % of the nation 65+ in age as predictors for national reported log(CFR). Lower WAIC values correspond to better model performance, and the reported weight is the proportional expanatory value of each model.

| Model | WAIC | dWAIC | Weight |
| --- | --- | --- | --- |
| $\rho + \text{GDP} + \text{Age}$ | 348.2 | 0.0 | 0.72 |
| $\rho + \text{GDP}$ | 351.0 | 2.8 | 0.18 |
| $\rho$ | 352.8 | 4.6 | 0.07 |
| $\rho + \text{Age}$ | 354.1 | 6.0 | 0.04 |

## Discussion

Estimating the infection fatality rate from the case fatality rate is not possible with limited testing capacity, though many policies regarding the COVID-19 pandemic rely on estimates of the severity of the disease. IFR is the primary metric upon which decisions have been based, and estimates range from 0.1% to over 3% [6, 8, 17, 18, 19]. The variety in reported CFR values stems from the variation in demography of study groups, income, access to healthcare, testing fidelity, or study design. Lack of large sample size or non-random selection of study participants hinders true estimates of the IFR in any locality. The estimate of IFR in one locality will differ from the IFR in another due to differences in underlying health conditions, demography, and medical treatment. Rather than understanding the extent of the variability these factors can create in IFR measurements, we derive a new approach to IFR estimation using global data. Provided testing has prioritized people at highest risk of SARS-COV-2 infection, the metric of relative testing capacity used in our approach should reflect each country’s ability to accurately estimate the true IFR. Despite variation in reporting and testing strategy, we do find a close relationship between testing capacity and reported IFR. Moreover, the relationship aligns well with the predicted relationship given assumptions regarding testing capacity. Transformation of the data using equation 9 results in a linear relationship shown in panel A of Figure 2, and the weighted regression provides a precise estimate of the global IFR of 1.04%. This estimate will vary by country, and the equations detailed in this study should not be used to estimate country level IFR without many independently derived samples of varying testing capacity as shown in this study. Equation 3 approximates global discovery of true positive cases, but it may not be reliable at smaller scales due to variation in testing protocol.

Deviation of reported CFR by each country from the global mean estimated from the regression line does not necessarily reflect true differences in national IFR from the global IFR. There is little to no correlation between these deviations and GDP or age, which are considered proxies of risk for mortality from COVID-19 [7, 21]. Instead, deviations likely represent variation in testing and reporting strategies. Table 1 gives the countries outside of the 90% confidence interval of the estimated global mean at their reported *ρ*. These countries likely vary from expectation due to testing differences that change estimates of testing capacity or differences in reporting deaths from COVID-19. Additional variation can occur because COVID-19 associated deaths lag diagnosis, so countries early in an epidemic may report a lower than expected CFR.

COVID-19 associated deaths may be positively identified through a PCR-based test or reported if medical diagnostic criteria are met as judged by a physician [20]. Due to comorbidities, such as hypertension or diabetes, estimates of COVID-19 deaths have been questioned. Arguments that death coinciding with a positive SARS-COV-2 test does not necessarily meet criteria for a COVID-19 associated death have been made. Given results of this work that indicate the risk of death to be approximately 1% from COVID-19, which is higher than the risk of death in the same 2-3 week interval from common chronic comorbidities [24, 25], the probability that a SARS-COV-2 positive death could be attributed to COVID-19 is higher than the probability the death could be attributed to other chronic comorbidities. In an estimate of 0.1% probability of mortality in a 2 week window due to existing comorbidities, the probability that death of an infected patient with a pre-existing condition is primarily due to the existing condition is less than 10% (SI 6). In this case, the reported CFR needs to be adjusted to 90% of its value. In light of this, the vast majority of SARS-COV-2 positive deaths are due in whole or primarily to COVID-19. High reported CFR values are a result of under-counting of positive infections rather than overcounting COVID-19 associated deaths. This contrasts needed adjustments in deaths associated with influenza, as the lower IFR of active influenza infections leads to a higher coincidence of mortality due to chronic disease and infection relative to the incidence of influenza associated mortality alone. In some localities it is estimated that COVID-19 associated deaths are under-reported by 25-50% because these deaths occur outside of a hospital setting [26]. Underreporting of deaths is likely to outweigh over-reporting due to comorbidities in many localities.

The inflation of reported CFR values can be approximated by testing capacity, as we relate in this study. The remaining 45% of the variation in the data unexplained by the metric of testing capacity used here is likely explained by a mixture of variability in national reporting, testing strategies, and stage of the epidemic with select cases explained by demography, underlying health conditions, income, or availability of treatment. The methods provided in this work and the supplement are easy to translate to other studies when high quality data and independent samples are available. We hope this approach can be used to give updated estimates of the global IFR, as deaths due to infection often lag diagnosis. Reported CFR values for some countries in this study are low due to the nascency of viral transmission in the country in April 2020. As more data becomes available for infections and COVID-19 deaths, case fatality rates can be more reliably approximated, which can be translated to a global IFR. Temporal or regional variation in testing strategy affects the relationship between CFR and testing capacity, therefore cumulative data collected at least 1 month after the first recorded COVID-19 death in each country better represents the relationship than point estimates. Best estimates rely on standardized global reporting practices, though this method is robust despite differences. Of note, some countries have yet to report deaths, and more have yet to report the number of tests administered, including China. Failure to report these estimates impairs resolution of the global IFR, which is crucial in providing a predictive framework for epidemiologists and policy makers to inform appropriate control measures.

The aim of this study was to provide a reliable metric that enables a robust estimate of the global IFR capturing a wide range of variation without accounting for underlying variability that is necessary to give point estimates of the IFR in each locality. Though we provide the first measurement, the estimate of a global IFR of 1.04% is dynamic and can be updated with incoming data. This number is lower than the IFR for the Diamond Princess cruise (2%), whose demography was skewed toward older individuals [18]. The older age of this sample may be partially counterbalanced by the higher income and access to healthcare of the majority of these individuals than the global average. National surveys performed in Iceland have estimated an IFR below 0.6%, which is among the most randomized testing conducted in any nation, though much of these cases have affected a younger group than is representative of the nation [27]. A previous estimate of IFR in China of 0.66% aligns well with projections from Iceland [19]. Seroprevalence studies have inferred widely different estimates of IFR, including a non-random sample of Santa Clara county residents in CA that suggested an IFR of 0.12-0.2% [17]. NYC seroprevalence of grocery shoppers suggests approximately one of every 5 NYC residents has been exposed. Because 0.15% of NYC residents had died from COVID-19 at the time the study was conducted, seroprevalence results yield a city-wide IFR of 0.75%. The variety of IFR estimates reflects study design and underlying risk factors that contribute to COVID-19 mortality. Although randomized seroprevalence surveys will provide the most accurate measurement of local IFR, these surveys cannot be conducted on a global scale, especially if the results of these surveys are to be used proactively to inform control strategies. Much of the current local estimates of IFR are representative of high income areas with high access to treatment. Higher IFR is possible in areas where surveys have not been conducted due to disparity in healthcare access or testing [21]. The IFR reported in this study attempts to capture some of the variability associated with these disparities between countries, which has otherwise been neglected in previous studies.

Most importantly, the realized IFR is dependent on healthcare capacity, which can vary with conditions of the pandemic. High case load can overwhelm hospital staff and resources, leading to high realized IFR. This is a possible explanation for the high departure of Italy’s reported CFR from the global estimate for equivalent testing capacity [28]. We estimate the IFR for Italy as 1.91% (SI 7), which is higher than a recent estimate of 1.29% provided by Rinaldi and Paradisi from early data but within their credible interval [29]. The IFR for Italy is likely higher than for countries of equivalent GDP and healthcare access due to the spike in cases that occurred in March 2020 that exhausted hospital resources in the northern regions. Proper control measures and testing can minimize infection spikes to levels that hospitals can manage, reducing the IFR.

## Data Availability

All data was collected from publicly available repositories.

https://www.worldbank.org/

https://www.worldometers.info/coronavirus/

https://github.com/pcm-dpc/COVID-19/tree/master/dati-regioni

## Contributors

REG conceived the study, extracted and analyzed data, developed the methods and code, and wrote the manuscript. GDL edited the manuscript and contributed to the analysis and final draft.

### Declaration of interests

Authors declare no competing interests.

### Data sharing

All data was collected from publicly available repositories: https://www.worldometers.info/coronavirus/,https://www.worldbank.org/, and https://github.com/pcm-dpc/COVID-19/tree/master/dati-regioni. P-values are available in the supplement, and code is available at [insert address].

## Acknowledgements

Many thanks go to the De Leo lab for support and to Kalin Wilson for additional comments on drafts of this manuscript. REG is funded by the Stanford Graduate Fellowship and Stanford Biology Department.

## Supplementary Information

## 1. Equation 3

Equation 3 in the main text can be derived from simple principles given a finite population of size *n*. In this population there is an underlying prevalence of COVID-19 infection such that this true prevalence 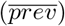 is represented by

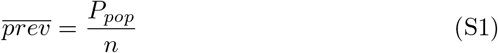

Discovery of the true number of cases, *P_pop_* in the finite population with increased sampling depends on the information available on the likelihood of infection with COVID-19. For a discrete process whereby sampling of the population for COVID-19 cases occurs iteratively, such that a defined subsample of the population is tested in each iteration, the proportion of the population sampled can be denoted *z*. Each subsample, *t*, changes the available prevalence of cases in the finite population when information is available to bias sampling in favor of testing COVID-19 positive individuals. This information exists because of the presence of symptoms or suspicion of exposure to infection. The bias associated with the information is denoted *λ* > 1. When *λ* = 1, sampling is random. Bias can be defined as the fold increase in likelihood of sampling an infected individual relative to sampling an uninfected individual. The prevalence of COVID-19 positive individuals in the unsampled population can be represented by *r_t_* where 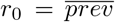 is the true prevalence (prior to sampling without replacement) in the population. Each iteration, *t*, is not by definition a time-dependent index. Rather, it is related to the number of tests conducted on the finite population, and is thus associated with sampling effort. When the population size is fixed, *t* can be indirectly related to time by way of the time taken to conduct *t* rounds of tests. The following relation can be found:

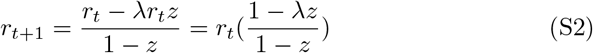

It follows that

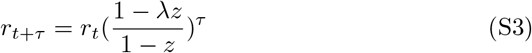

Therefore, the following relation is true:

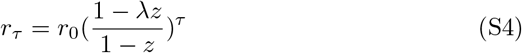

The sample prevalence can be represented as

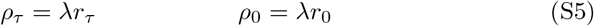

Substitution yields

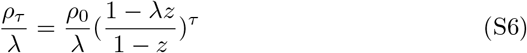

Rearrangement gives

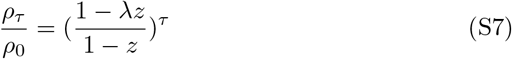

The total number of positive test results achieved can be calculated as

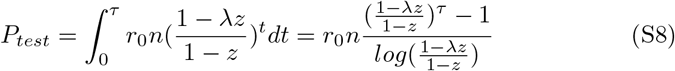

From equation S1, the number of cases in the population is

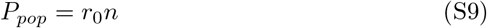

Combining equations S8 and S9 yields

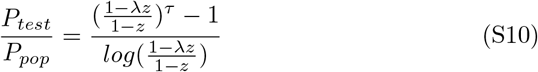

From equation S7 we find

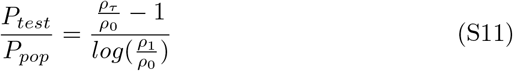

As *ρ*_0_ is dependent on the true prevalence in the population and is the maximum in the set {*ρ_t_*}, the sample prevalence at *τ* can be adjusted to a reported sample prevalence *ρ* that can vary between 0 and 1 rather than 0 and *ρ*_0_.

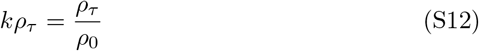

Denoting Δ as the quotient of *ρ*_1_ and *ρ*_0_ gives a simplified equation

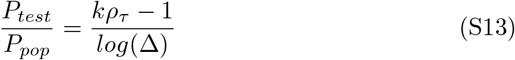

For *P_test_* to converge to *P_pop_* as *ρ*_τ_ → 0, Δ has a unique real valued solution at *e*^-1^. The equation becomes

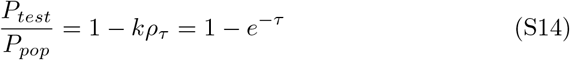

The Taylor expansion of 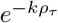 at *ρ_τ_* = 0 is 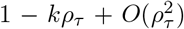, therefore supplemental equation 14 can be approximated as

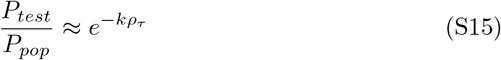

Equation S14 is an exact relation that represents a closed system in which sampling occurs much more quickly than the accumulation of new infections. Even under slow sampling, convergence to the true population case load occurs at a constant rate. However, when new cases occur at a similar rate to testing, convergence should occur more slowly. Equation S15 has the capacity to capture this property more accurately at high *ρ_τ_* (slow sampling) than equation S14 while maintaining the same properties at low *ρ_τ_* (high sampling).

Equation 3 in the main text reports the relationship between testing capacity and the convergence of the positively identified COVID-19 cases and the true number of cases in the population, which is the sum of the identified and unidentified active cases. This relationship will differ with testing regime, as in each locality procedures for administering tests vary, including the frequency of re-testing that occurs. However, the function should approach 1 as *C → ∞*. With a higher proportion of symptomatic patients, more information is available to prioritize testing, so convergence of the estimate of cases occurs more rapidly with increasing capacity. 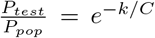 provides a simple function with the desired property of approaching 1 as *C → ∞*. Use of this equation provides the derivation of equation 9 in the main text, which gives the relationship between CFR and C. Transforming the data by inverting the x-axis and log-transforming the y-axis produces a linear relationship as predicted by equation 9. In this way, the use of equation 3 is supported, though because it is not otherwise empirically verified, its use should not be extended to approximate local case loads. As mentioned in the main text and here, variation in testing protocol invalidate use of equation 3 at small scales.

### 1.1. Parameter de_nitions

**Table 3:** List of parameters in order of appearance. The dashed line separates parameters defined in the main text from those defined in the supplement.

| Parameter | Definition |
| --- | --- |
| $C$ | Testing capacity |
| $T$ | No. SARS-COV-2 tests conducted in country |
| $P_{test}$ | No. positive test results in country |
| $\rho$ | Sample prevalence at given testing effort $\tau$ (same as $\rho_\tau$) |
| $P_{pop}$ | No. cases in country at a given time |
| $k$ | Slope of linear regression and decay constant |
| $CFR$ | No. of deaths per positive test result |
| $IFR$ | No. of deaths per infection |
| $N$ | No. of countries |
| $\overline{prev} = r_0$ | Infection prevalence in population |
| $n$ | Size of population |
| $z$ | Proportion of population tested per iteration $t$ |
| $t$ | Index of testing effort |
| $\tau$ | Total testing effort |
| $\lambda$ | Sampling bias in favor of infected individuals |
| $r_t$ | Infection prevalence after $\tau$ testing effort |
| $\Delta$ | $\rho_1/\rho_0$ |
| $\sigma_i^2$ | Sample variance for country $i$ |

## 2. Linear regression

The linear regression reported in Figure 2 (main text) was performed according to equation 10 with inverse variance weights. Variance in reported CFR values for each country is a function of the sample variance 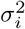 due to testing protocol, specificity, and accuracy, and the number of tests performed *n_i_*.

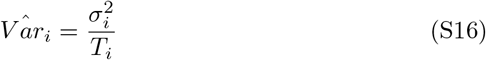

Because sample variance has not been measured for each country, we assume {*σ^2^*} is homoscedastic across countries. The estimate of variance is therefore inversely proportional to the number of tests performed by each country. Weights are then assigned as

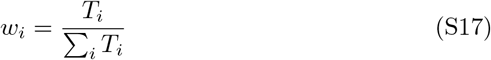

These weights are used as the loss function on the squared residuals. Equation 10 (main text) is minimized with a ‘BFGS’ optimization algorithm provided by the Python scipy.optimize.minimize routine. Parameters *log*(*ÎFR*) and 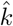 are estimated through minimization of the weighted residuals, and the Hessian is numerically estimated and inverted. This provides a scaled variance-covariance matrix that is multiplied by the *RSS* and divided by the degrees of freedom to give the variance-covariance matrix, **V** (equation 11 main text). The standard error of each parameter is

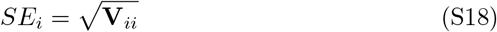

The covariance of 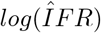 and 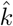 is reported in the two off-diagonal entries of **V**.

**Figure S1:**
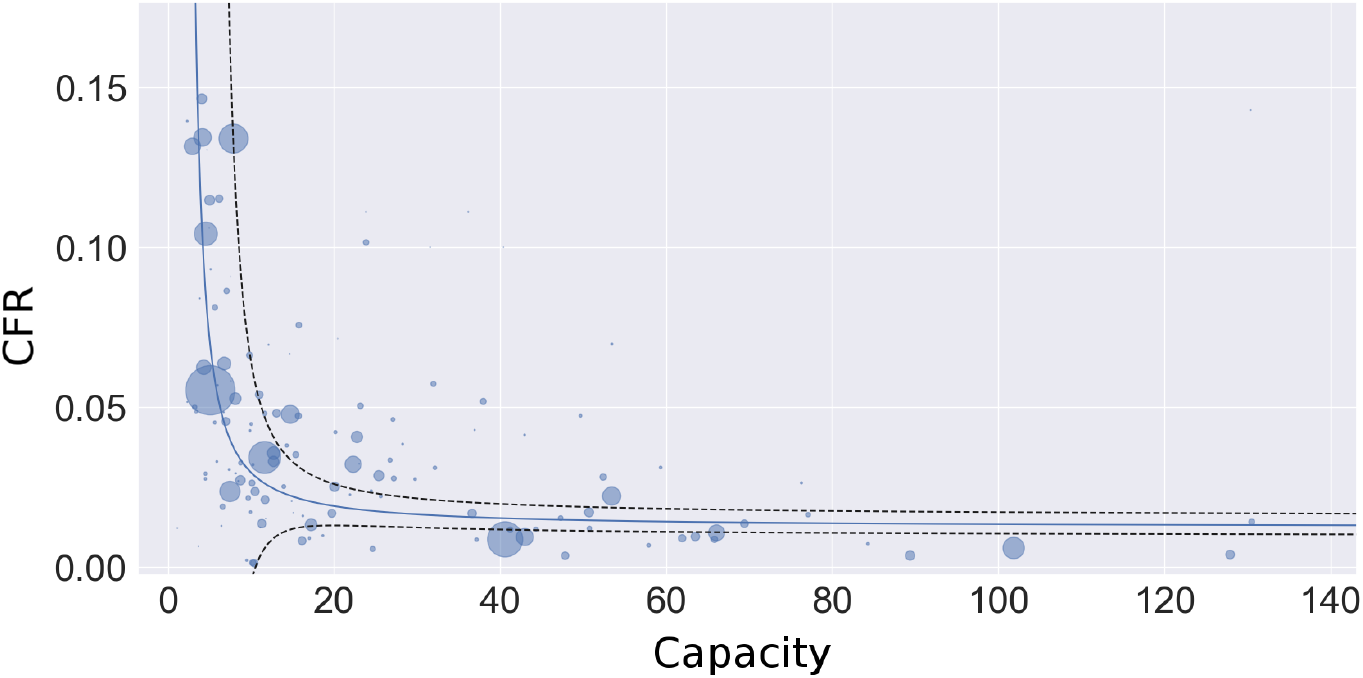
Plot of raw data of capacity vs CFR by country with superimposed estimated regression. In contrast to Figure 2 in the main text, the capacity axis is not log-scale. Venezuela is not plotted due to its high capacity.

## 3. *R*^2^ calculation

The *R^2^* reported from the linear regression in Figure 2 panel A of the main text is given as follows:

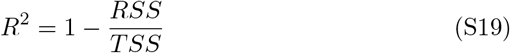

RSS is defined as:

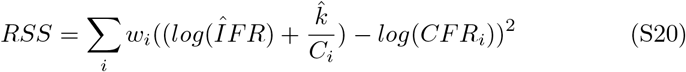

TSS is defined as:

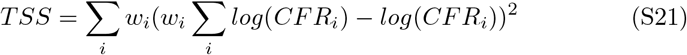

## 4. Confidence interval estimation

The linear regression on the log-transformed data provides a estimated line with normally distributed errors, and the 95% confidence intervals for 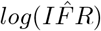 is easily approximated as 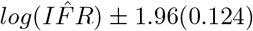, where 0.124 is the standarderror of the estimated parameter. The same calculation provides a 95% confidence interval for 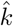, approximated as 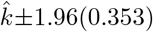, where 0.353 is the standard error for 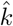. A confidence interval for 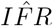 requires exponentiating 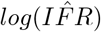 to produce a log-normal distribution. Letting 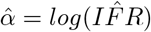, the confidence interval becomes 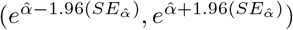. This interval can be used to propagate error in the nonlinear regression. To do so, we define values which represent the standard difference between the estimate and the 95% bounds:

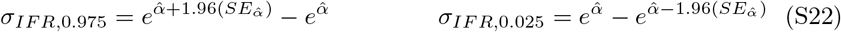

The 95% confidence intervals for the linear regression can be calculated as

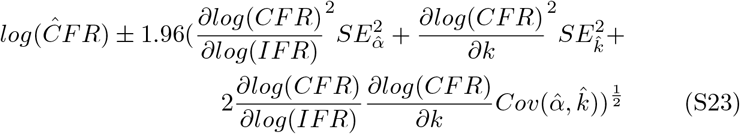

Partial derivatives are calculated as

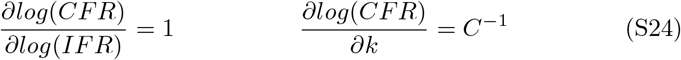

Therefore, the confidence interval simplifies to

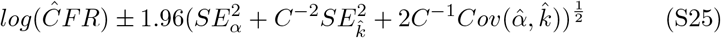

These intervals are reported as the dashed lines in Figure 2 panel A (main text). Nonlinear 95% confidence intervals are estimated using the values defined in equation S25. Upper and lower bounds are described as

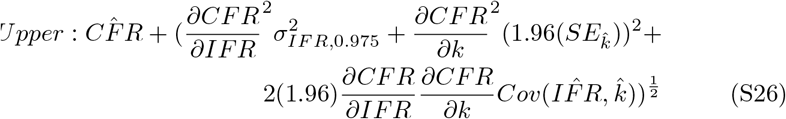

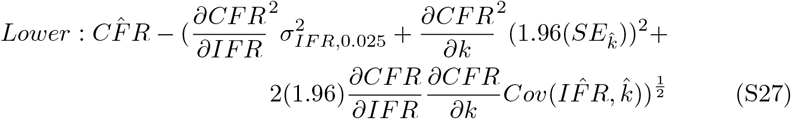

Partial derivatives are calculated as

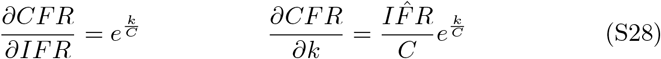

Therefore, the confidence interval simplifies to

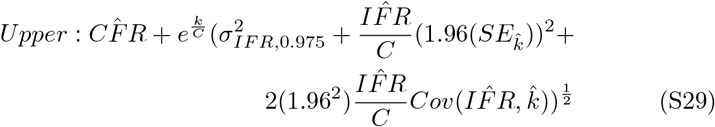

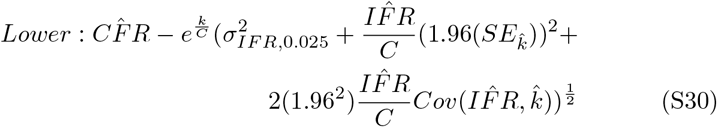

The covariance of the parameters in the linear regression, 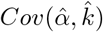, is given by the Hessian-derived variance-covariance matrix, **V**. However, because of the exponentiation of *α* to derive 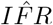, the covariance between the parameters no longer applies. 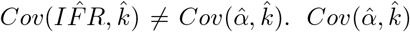 requires derivation. Let ξ be the log-normally distributed random variable representing *IFR* with median at 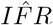. Let *α* be the normally distributed random variable representing *log*(*IFR*) with mean at 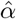. *k* is a normally distributed random variable with mean at 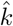. *ξ* = *e^α^*

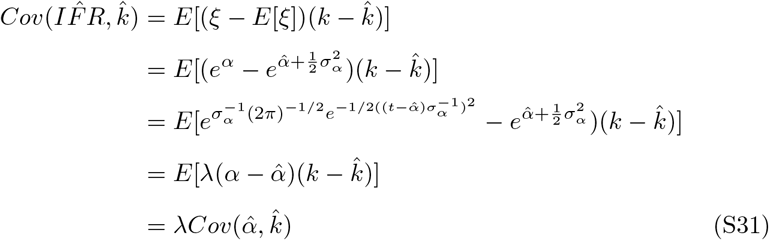

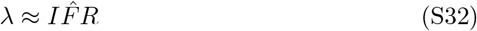

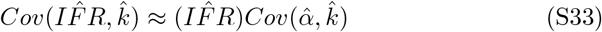

This approximation was verified by monte carlo simulation at 10^7^ replicates.

## 5. Statistical test for deviation

A Welch’s one-way T-test for unequal variances was conducted to determine the significance of the deviation of national CFR values from the expected regression estimate. The mean and standard error for the regression estimate are available for each value of *ρ* = *C*^−1^ corresponding to each nation’s data. The standard error for reported national CFR values was calibrated to the confidence interval reported in the regression such that 95% of p-values fell within 0.025 *< p <* 0.975. Calibration occurred assuming homoscedasticity in the sampling variance, and denominator of the variance estimate was given as the number of tests administered by each country. The p-values were used in the linear regressions of Figure 3 in the main text. Extreme p-values were reported in Table 1 of the main text. Table 1 of the supplement gives the full list of p-values by country.

**Table 4:**
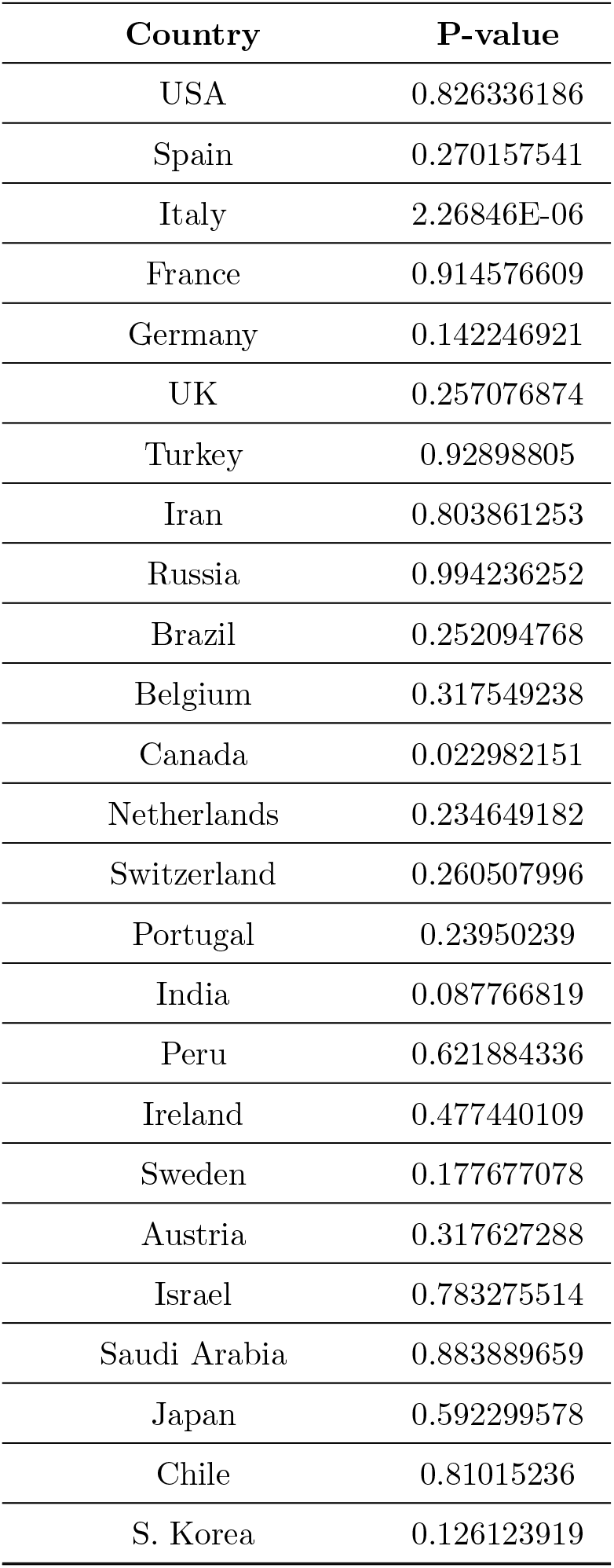
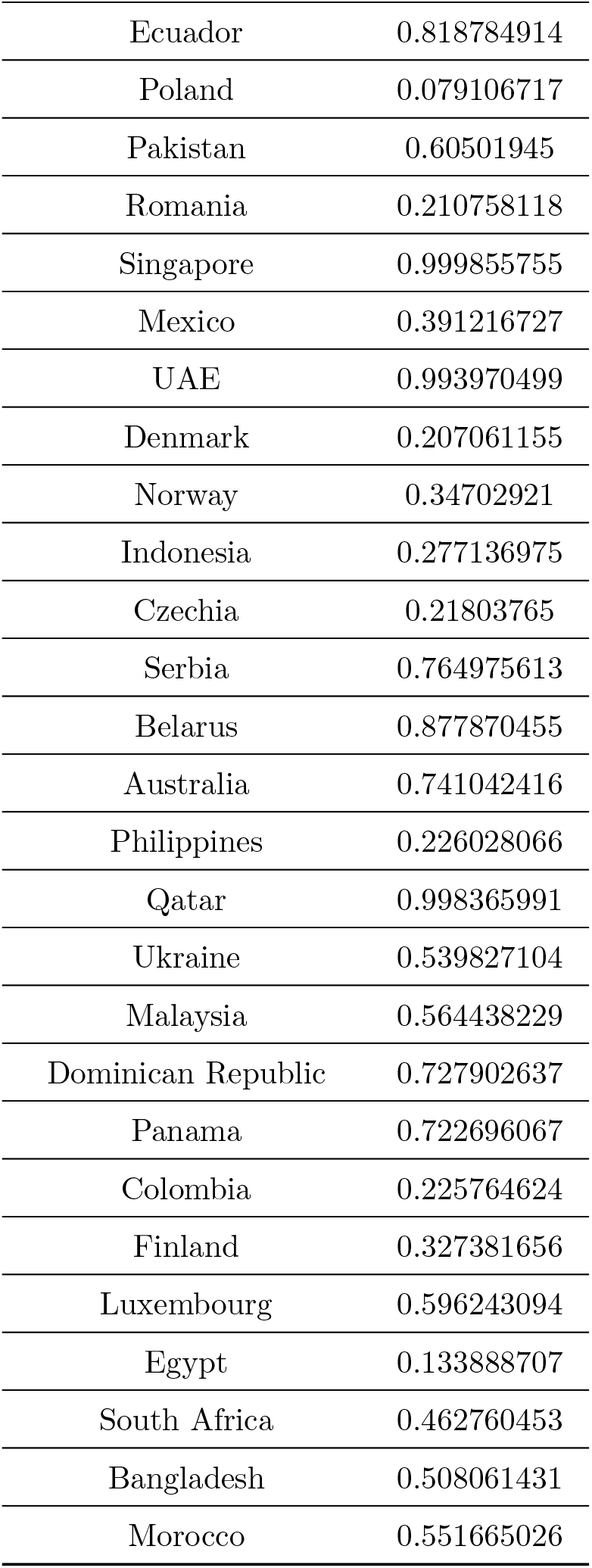
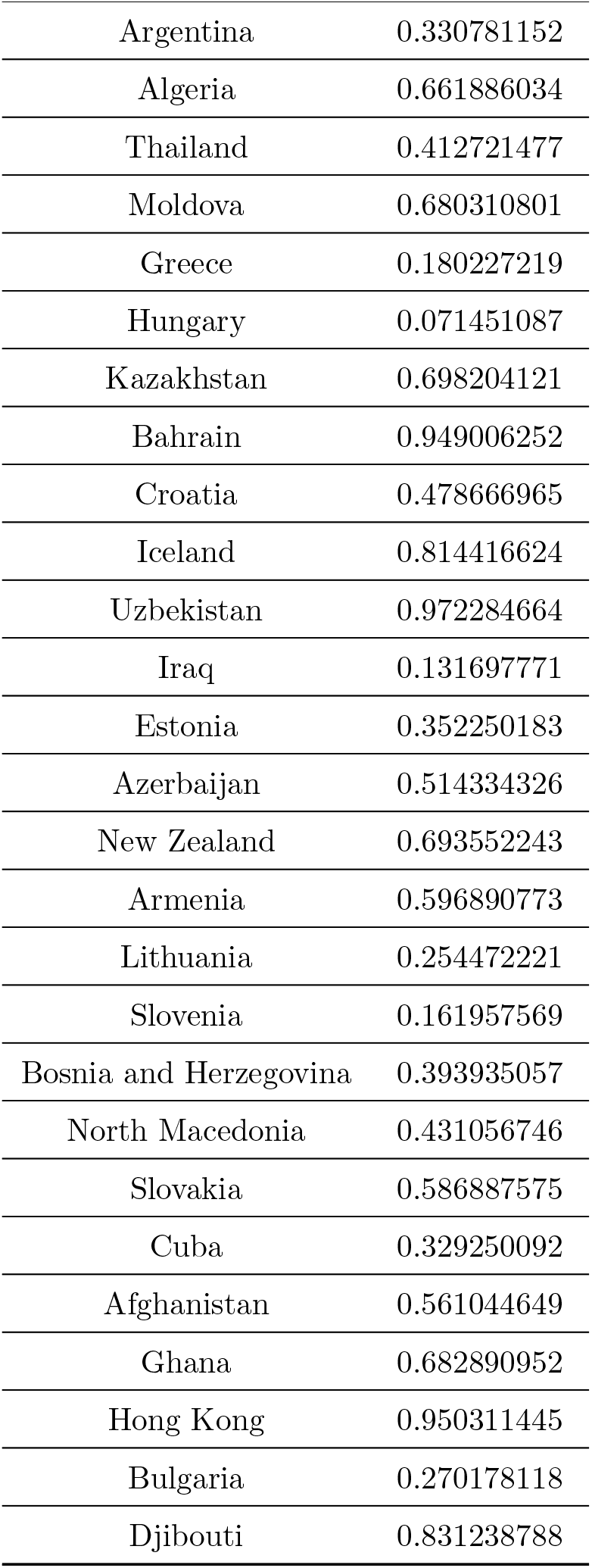
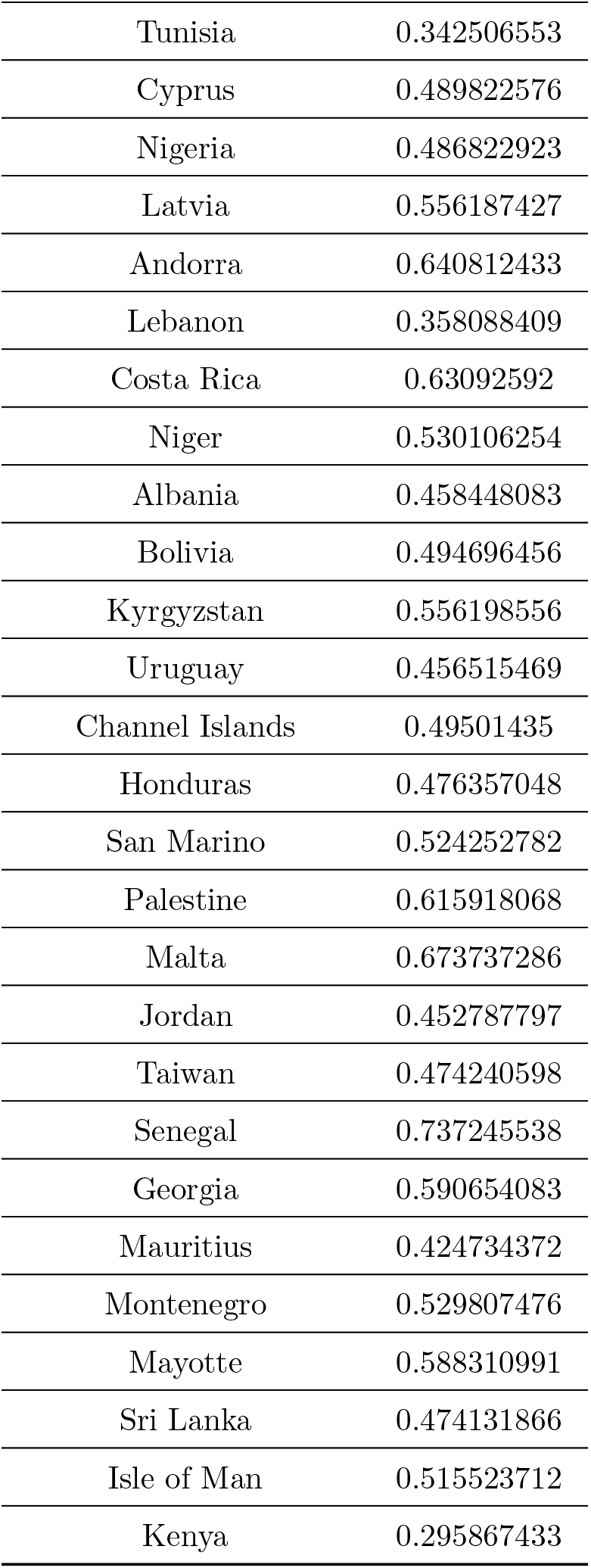
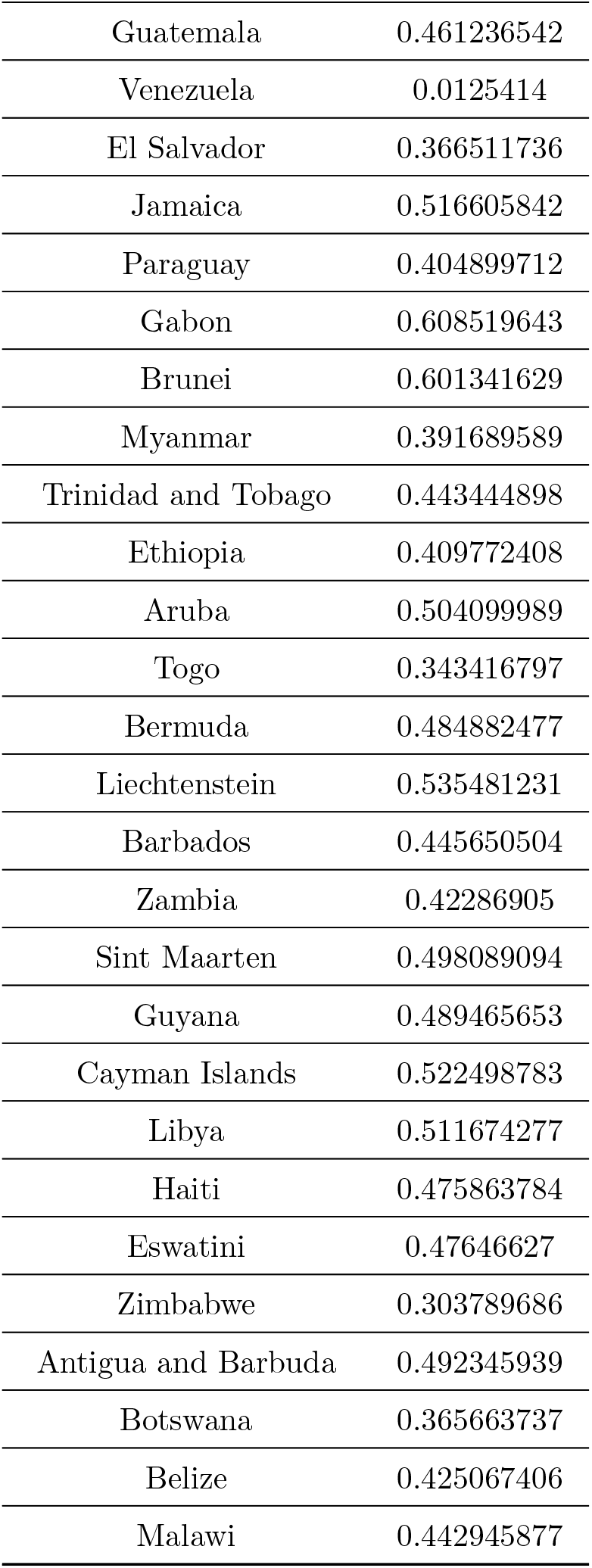
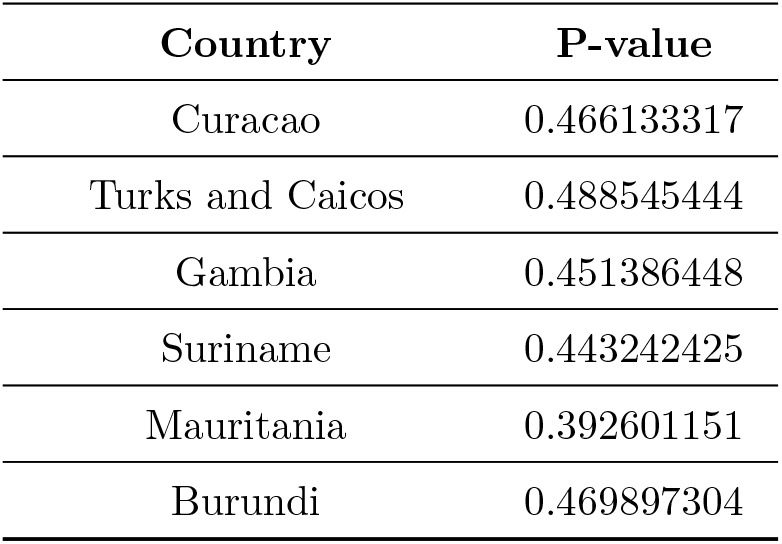
P-values for deviation analysis. Countries are ordered by caseload as of April 21, 2020. Higher p-values indicate lower than expected reported CFR. Lower p-values indicate higher than expected reported CFR.

| Country | P-value |
| --- | --- |
| USA | 0.826336186 |
| Spain | 0.270157541 |
| Italy | 2.26846E-06 |
| France | 0.914576609 |
| Germany | 0.142246921 |
| UK | 0.257076874 |
| Turkey | 0.92898805 |
| Iran | 0.803861253 |
| Russia | 0.994236252 |
| Brazil | 0.252094768 |
| Belgium | 0.317549238 |
| Canada | 0.022982151 |
| Netherlands | 0.234649182 |
| Switzerland | 0.260507996 |
| Portugal | 0.23950239 |
| India | 0.087766819 |
| Peru | 0.621884336 |
| Ireland | 0.477440109 |
| Sweden | 0.177677078 |
| Austria | 0.317627288 |
| Israel | 0.783275514 |
| Saudi Arabia | 0.883889659 |
| Japan | 0.592299578 |
| Chile | 0.81015236 |
| S. Korea | 0.126123919 |

| **Country** | **P-value** |
| --- | --- |
| Ecuador | 0.818784914 |
| Poland | 0.079106717 |
| Pakistan | 0.60501945 |
| Romania | 0.210758118 |
| Singapore | 0.999855755 |
| Mexico | 0.391216727 |
| UAE | 0.993970499 |
| Denmark | 0.207061155 |
| Norway | 0.34702921 |
| Indonesia | 0.277136975 |
| Czechia | 0.21803765 |
| Serbia | 0.764975613 |
| Belarus | 0.877870455 |
| Australia | 0.741042416 |
| Philippines | 0.226028066 |
| Qatar | 0.998365991 |
| Ukraine | 0.539827104 |
| Malaysia | 0.564438229 |
| Dominican Republic | 0.727902637 |
| Panama | 0.722696067 |
| Colombia | 0.225764624 |
| Finland | 0.327381656 |
| Luxembourg | 0.596243094 |
| Egypt | 0.133888707 |
| South Africa | 0.462760453 |
| Bangladesh | 0.508061431 |
| Morocco | 0.551665026 |

| **Country** | **P-value** |
| --- | --- |
| Argentina | 0.330781152 |
| Algeria | 0.661886034 |
| Thailand | 0.412721477 |
| Moldova | 0.680310801 |
| Greece | 0.180227219 |
| Hungary | 0.071451087 |
| Kazakhstan | 0.698204121 |
| Bahrain | 0.949006252 |
| Croatia | 0.478666965 |
| Iceland | 0.814416624 |
| Uzbekistan | 0.972284664 |
| Iraq | 0.131697771 |
| Estonia | 0.352250183 |
| Azerbaijan | 0.514334326 |
| New Zealand | 0.693552243 |
| Armenia | 0.596890773 |
| Lithuania | 0.254472221 |
| Slovenia | 0.161957569 |
| Bosnia and Herzegovina | 0.393935057 |
| North Macedonia | 0.431056746 |
| Slovakia | 0.586887575 |
| Cuba | 0.329250092 |
| Afghanistan | 0.561044649 |
| Ghana | 0.682890952 |
| Hong Kong | 0.950311445 |
| Bulgaria | 0.270178118 |
| Djibouti | 0.831238788 |

| **Country** | **P-value** |
| --- | --- |
| Tunisia | 0.342506553 |
| Cyprus | 0.489822576 |
| Nigeria | 0.486822923 |
| Latvia | 0.556187427 |
| Andorra | 0.640812433 |
| Lebanon | 0.358088409 |
| Costa Rica | 0.63092592 |
| Niger | 0.530106254 |
| Albania | 0.458448083 |
| Bolivia | 0.494696456 |
| Kyrgyzstan | 0.556198556 |
| Uruguay | 0.456515469 |
| Channel Islands | 0.49501435 |
| Honduras | 0.476357048 |
| San Marino | 0.524252782 |
| Palestine | 0.615918068 |
| Malta | 0.673737286 |
| Jordan | 0.452787797 |
| Taiwan | 0.474240598 |
| Senegal | 0.737245538 |
| Georgia | 0.590654083 |
| Mauritius | 0.424734372 |
| Montenegro | 0.529807476 |
| Mayotte | 0.588310991 |
| Sri Lanka | 0.474131866 |
| Isle of Man | 0.515523712 |
| Kenya | 0.295867433 |

| **Country** | **P-value** |
| --- | --- |
| Guatemala | 0.461236542 |
| Venezuela | 0.0125414 |
| El Salvador | 0.366511736 |
| Jamaica | 0.516605842 |
| Paraguay | 0.404899712 |
| Gabon | 0.608519643 |
| Brunei | 0.601341629 |
| Myanmar | 0.391689589 |
| Trinidad and Tobago | 0.443444898 |
| Ethiopia | 0.409772408 |
| Aruba | 0.504099989 |
| Togo | 0.343416797 |
| Bermuda | 0.484882477 |
| Liechtenstein | 0.535481231 |
| Barbados | 0.445650504 |
| Zambia | 0.42286905 |
| Sint Maarten | 0.498089094 |
| Guyana | 0.489465653 |
| Cayman Islands | 0.522498783 |
| Libya | 0.511674277 |
| Haiti | 0.475863784 |
| Eswatini | 0.47646627 |
| Zimbabwe | 0.303789686 |
| Antigua and Barbuda | 0.492345939 |
| Botswana | 0.365663737 |
| Belize | 0.425067406 |
| Malawi | 0.442945877 |
| Curacao | 0.466133317 |
| Turks and Caicos | 0.488545444 |
| Gambia | 0.451386448 |
| Suriname | 0.443242425 |
| Mauritania | 0.392601151 |
| Burundi | 0.469897304 |

## 6. Comorbidities and assignment of death to COVID-19

Estimating the mortality due to COVID-19 is not clear for many cases because infection can co-occur pre-existing conditions that put patients at risk of death upon infection. There is an argument to be made for accurately assigning cause of death, and infection with COVID-19 may not be sufficient. Analogies have been made to estimates of mortality due to influenza, which are notoriously variable depending on the criteria used to assign mortality. The vast majority of patients that die from influenza have pre-existing conditions, much like those that die from COVID-19. Therefore, many influenza associated deaths could be attributed to pre-existing conditions. Although for both influenza and COVID-19 infections, pre-existing conditions may be associated with higher risk of mortality, there are major differences in the responsibility comorbidites share in death between these infectious diseases. The primary difference is due to the greater IFR for COVID-19 and influenza. For symptomatic infections, the CFR for influenza is 0.1%. Accounting for asymptomatic patients, which may be between 15 and 85% of cases (we will assume a conservative estimate of 25%), the IFR is approximately 0.00075. The estimated IFR for COVID-19 from this study is approximately 0.01. There is mounting evidence suggesting overlapping comorbidities between influenza and COVID-19. Assuming the window of mor-tality due to infection from influenza or COVID-19 is approximately 2 weeks, the probability of death from one of the most likely comorbidities like heart disease or diabetes in the two week window is approx. 10^−3^ (after diagnosis and 50+ yrs of age, USA) [30]. The harmonic mean of the life expectancy for men (6.7 yrs) and women (7.9 yrs) over 50 diagnosed with heart disease in the USA is 7.25 yrs. The probability of death in a two week window assuming constant death rate is

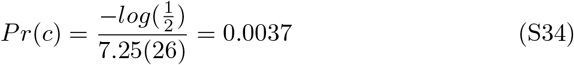

Similar calculations for diabetes yields *Pr*(*c*) < .001. Other pre-existing conditions associated with COVID-19 deaths have similar or lower death rates. Assuming contracting an infectious disease is independent of pre-existing conditions, the probability that the death of a patient with both a pre-existing condition and infection can be assigned primarily to the infection can be calculated. Given a death of a patient with a pre-existing condition (c = death by condition) and positive infection (i = death by infection), the probability the infection can be assigned the primary contributor to death is:

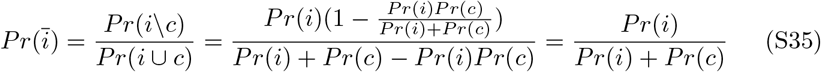

Given *Pr*(*c*) = 0.001 and *Pr*(*i*) = *IFR*, the probability an influenza associated death is due to influenza is:

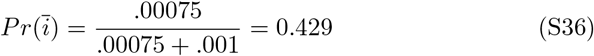

The probability a COVID-19 associate death is due to COVID-19 is:

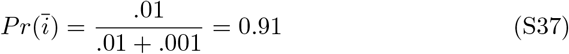

Although these calculations are simplifications of a more complex process (e.g. death rates are not constant with age), the magnitude of the IFR has a dramatic influence on the adjustment needed to assignment of cause of death. Adjustments required for estimates of an IFR for COVID-19 are far smaller than the adjustments needed for influenza. Likely, these adjustments will be less than 10% with an IFR above 1%. Lower adjustments are needed with fewer cases with comorbidities, and higher adjustments are needed when more than one comorbidity affect an individual. Assuming a 10% reduction in COVID-19 associated deaths would give a global IFR of 0.9%.

## 7. Timing of regression estimation

The regression estimate provided in the main text does not explicitly account for temporal variation in testing strategies or in CFR estimates for each country. Because these factors could have a strong impact on a country’s relationship between CFR and testing capacity, the best estimates of the relationship come near the peak of the epidemic in each country. This gives an opportunity for the estimate of the CFR in the country to level, as deaths lag diagnoses, and minimize subsequent swings in the relationship between testing capacity and accumulation of new cases. Figure S2 shows how the relationship between CFR and capacity can change over time with cumulative data for Italy. Using cumulative data from any day in March would yield similar results due to the linear nature of the relationship between sample prevalence and log(CFR) in this period.

Italy illustrates this concept well due to the easily measured peak in the epidemic near the end of March. Once deaths began to accumulate in each region in the beginning of March, the relationship between log(CFR) and *ρ* is linear until the peak of the epidemic where testing capacity outpaced new cases, and the number of positive cases per test drastically fell. Because cumulative data from Italy used in the global analysis presented in the main text was taken on April 21, 2020, this may explain the deviation of Italy from the expected curve. 4 weeks past the peak, data for Italy at the end of April is sub-optimal and predicts a higher CFR relative to capacity compared to countries that have yet to peak. April 21, 2020 remains a good choice for a single date measurement because many countries are near the peak or before the peak in the epidemic. Future estimates should focus on obtaining CFR and capacity measurements of cumulative data at the peak of each country’s epidemic. Best estimates also occur if testing capacity, as defined in the main text, is approximately constant over time. This indicates that the number of tests available keeps pace with growth and waning of case loads in the country. Variation in this relationship will not dramatically impact results, as cumulative data approximates the average over time. However, dramatic swings in testing capacity or testing protocol may impact reliability of estimates. Particular to this study, by April 21, most countries with reported deaths had experienced a few weeks or months of the pandemic. For these countries, estimates of the relationship between CFR and capacity are more reliable, particularly for the countries for which peaks in daily mortality had occurred. Several countries had few reported deaths and a growing case load due to later introduction of COVID-19 to the country or differences in control strategies. For these countries, estimates as applicable in a global regression will improve over time. The result of this approach can be updated to account for better estimates as the pandemic continues, though the global IFR is unlikely to fall far outside of the 95% confidence interval reported here. Italy provides a good example of the use of the approach developed in this study, as the epidemic growth occurred quickly, such that a peak is easily defined, and a range of dates near the peak is optimal for obtaining cumulative data and performing a regression. Doing so for Italy (at March 23, 2020), produces Figure S3.

**Figure S2:**
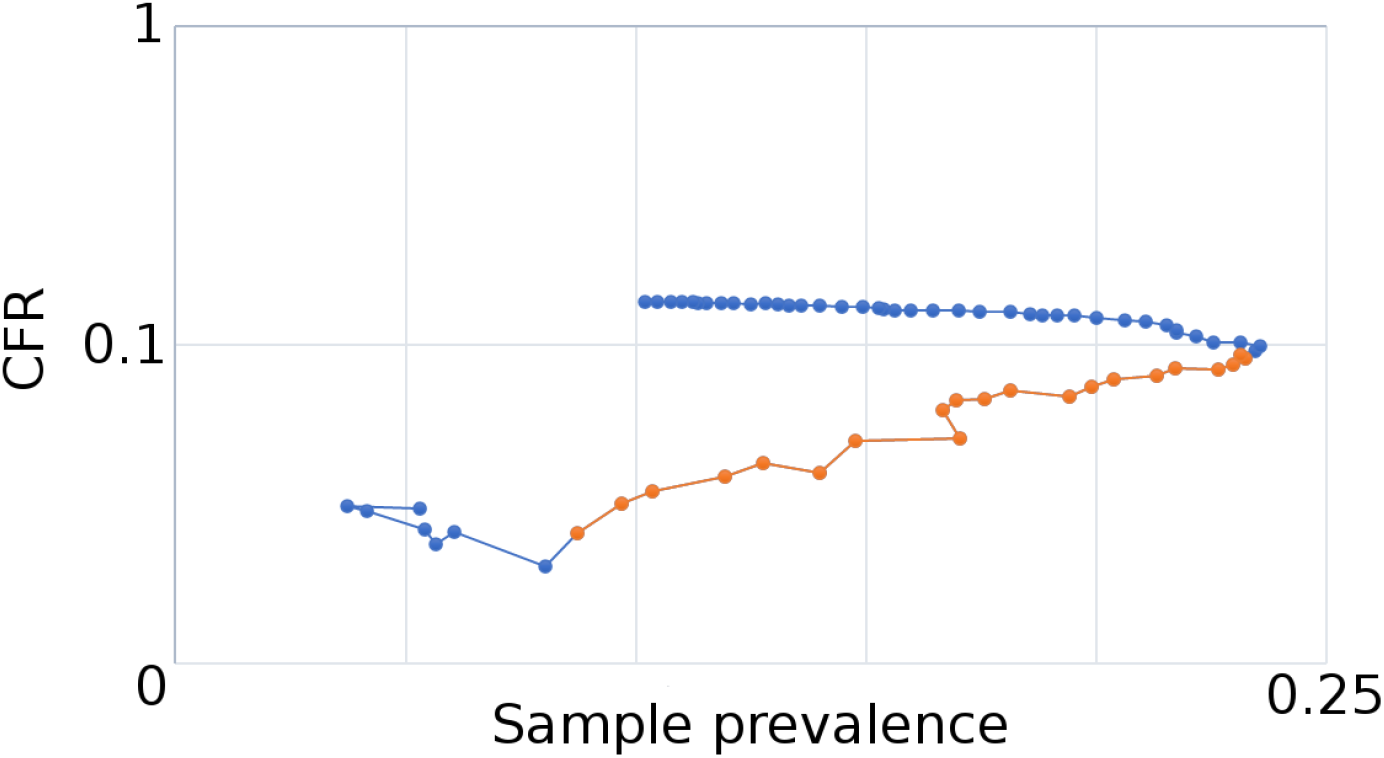
The relationship between CFR and capacity changes through the course of the epidemic in Italy. Orange points represent the rise in the epidemic (last orange point = March 22, 2020), demonstrating a close relationship between the scaling of testing and the rise in cases and fatalities. When new infections subside, testing capacity outpaces the accumulation of new cases relative to the previous weeks, changing the relationship. The estimator works best near the peak of the epidemic. Y-axis is reported as log-scale.

For some countries, such as the USA, a peak is not clearly defined due to regional heterogeneity in the course of the epidemic and variation in control strategies. Adjustment for false negatives in testing results in an adjusted IFR estimate for Italy at 1.91% (CI: 1.25%, 2.85 %). This estimate is almost twice as high as the global estimate, which is not unexpected given the high proportion of older individuals and the spike in cases that overwhelmed the healthcare system in Italy in March 2020.

**Figure S3:**
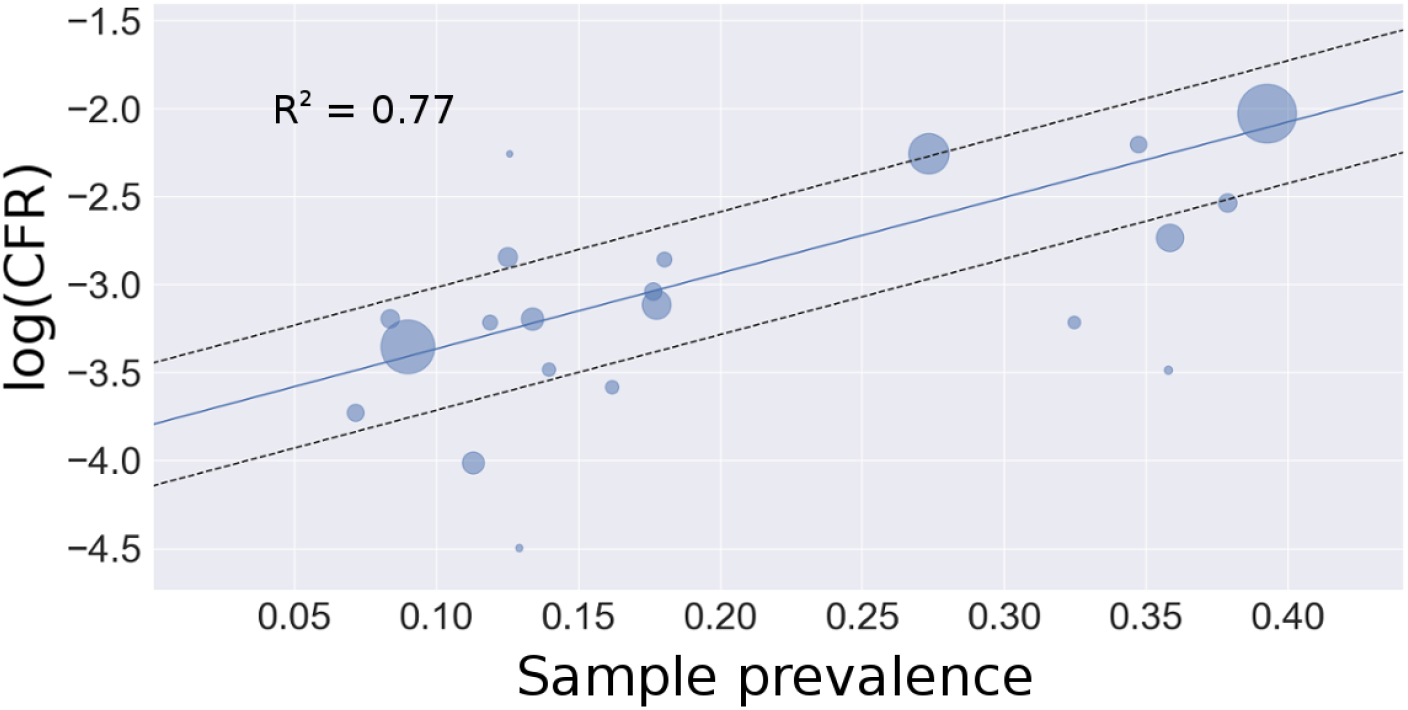
Regression estimate of Italy’s IFR using 20 Italian regions. The intercept corresponds to an unadjusted IFR of 2.24% (CI: 1.58%, 3.18 %).

## 8. Bayesian model selection

The generalized linear mixed model run to determine whether national per capita GDP or age (% 65+) added predictive value to the relationship between *ρ* and cumulative national *log*(*CFR*) revealed marginal improvement in the model fit as evaluated by AIC. The best fitting model included all three variables (*ρ*, per capita GDP, and age). 4 models were compared: *ρ* alone, *ρ* + per capita GDP, *ρ* + age, and all 3. Here we report the estimates provided by these models as well as raw output to demonstrate model convergence.

Model convergence is shown below

**Figure S4:**
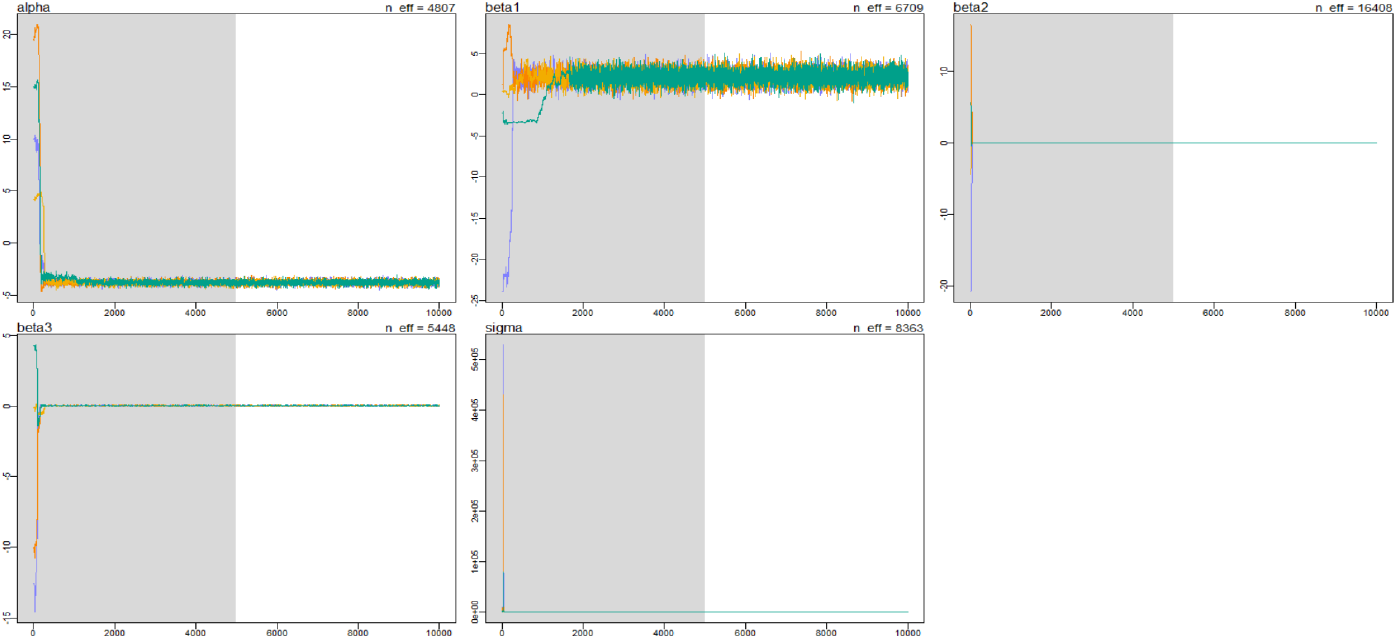
Plots of MCMC iterations, showing convergence for all parameter estimates. The shaded region gives the warmup period. *α =* intercept, *β*_1_ = *ρ*, *β*_2_ = GDP, *β_3_* = age, *σ* = random error of model. Colors represent each of 4 chains.

|  | mean | se_mean | sd | 2.5% | 25% | 50% | 75% | 97.5% | n_eff | Rhat |
| --- | --- | --- | --- | --- | --- | --- | --- | --- | --- | --- |
| alpha | -3.80 | 0.00 | 0.18 | -4.15 | -3.92 | -3.80 | -3.69 | -3.46 | 4807 | 1 |
| beta1 | 2.14 | 0.01 | 0.78 | 0.63 | 1.61 | 2.13 | 2.66 | 3.68 | 6709 | 1 |
| beta2 | 0.00 | 0.00 | 0.00 | 0.00 | 0.00 | 0.00 | 0.00 | 0.00 | 16408 | 1 |
| beta3 | 0.03 | 0.00 | 0.01 | 0.00 | 0.02 | 0.03 | 0.04 | 0.06 | 5448 | 1 |
| sigma | 0.93 | 0.00 | 0.06 | 0.82 | 0.89 | 0.92 | 0.97 | 1.05 | 8363 | 1 |
| dev | 337.76 | 0.04 | 3.23 | 333.55 | 335.42 | 337.09 | 339.37 | 345.75 | 6510 | 1 |
| lp__ | -53.89 | 0.02 | 1.62 | -57.90 | -54.69 | -53.55 | -52.72 | -51.78 | 6495 | 1 |

Raw output for model parameters.

The relationships among the model parameters are reported below in Figure S5.

**Figure S5:**
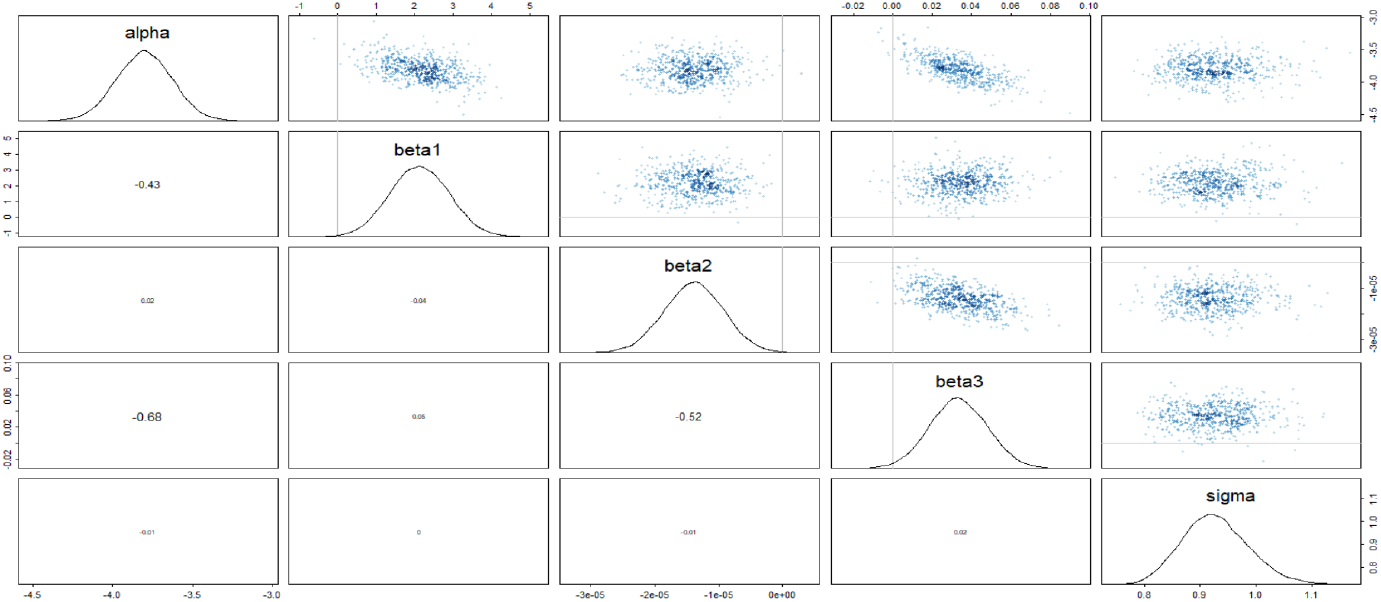
Distributions and relationships reported among model parameters. 2D density plots are given in the upper right triangle, while correlation coefficients are available in the corresponding bottom triangle. The diagonal reports the estimated distribution of each parameter and provides column and row labels. *α* = intercept, *β*_1_ = *ρ* coefficient, *β*_2_ = GDP coefficient, *β*_3_ = age coefficient, *σ* = random error of model.

The best performing model is specified as

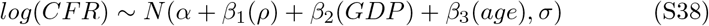

Priors were set as the following:

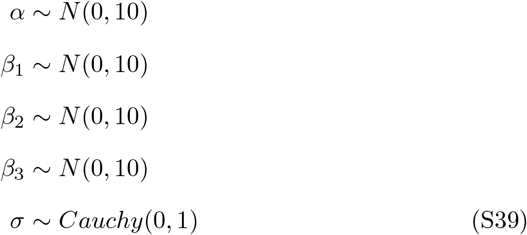

The estimate of *α* provides the intercept, which markedly differs from the weighted regression estimate of –4.4 provided in the main text. The difference here is primarily attributed to the lack of weighting in the Bayesian GLMM approach to compare variables. A weighting scheme was not appropriate for GDP and age data, and to compare *ρ* with the same framework to recover underlying relationships, this data was also unweighted. Additional differences can be attributed to residual influences of the other variables on the intercept.

## Notes

### Competing Interest Statement

The authors have declared no competing interest.

